# Thread Embedding Acupuncture for Temporomandibular Disorder: Protocol for a Randomized controlled Clinical Trial

**DOI:** 10.1101/2025.03.05.25323430

**Authors:** Jinkyung Park, Jung-Hyun Kim, Jaeho Song, Chanju Im, Jun-Gyu Park, Sang-Soo Nam, Bonhyuk Goo

**Affiliations:** Department of Acupuncture & Moxibustion, Kyung Hee University Hospital at Gangdong, Seoul, Republic of Korea; Department of Clinical Korean Medicine, Graduate School, Kyung Hee University, Seoul, Republic of Korea; Department of Acupuncture & Moxibustion, Kyung Hee University College of Korean Medicine, Kyung Hee University, Seoul, Korea

**Keywords:** Temporomandibular Joint Disorder, chronic pain, vertical mouth opening, Jaw Functional Limitation Scale, polydioxanone thread, acupuncture, DC/TMD

## Abstract

**Introduction:** Temporomandibular joint (TMJ) disorders are associated with severe pain and functional impairment. Although thread embedding acupuncture (TEA) is commonly used for the management of temporomandibular disorders (TMD) at clinical sites, evidence supporting its use remains insufficient. This study aimed to evaluate the efficacy, safety, and cost-effectiveness of TEA for the treatment of chronic TMJ disorders.

**Methods:** This randomized, assessor-blinded, controlled clinical trial included 76 patients with chronic TMD. Participants will be randomly allocated to either the TEA or the usual care group. The TEA group will receive treatment administered at eight bilateral predefined acupoints for six weeks. The usual care group will receive transcutaneous electrical nerve stimulation (TENS) and infrared therapy. The primary outcome was average pain intensity measured using the Visual Analog Scale (VAS). The secondary outcomes will include maximum pain intensity (VAS), vertical TMJ opening, Graded Chronic Pain 2.0, Jaw Functional Limitation Scale-20, Patient Global Impression of Change, EuroQol 5-Dimension 5-level (EQ-5D-5L), adverse events, and medical costs for economic evaluation.

**Discussion:** This study provides evidence for the efficacy and safety of TEA for chronic TMJ disorders and explores its potential as a cost-effective treatment.

**Trial registration:** This study was registered with the Clinical Research Information Service of the Republic of Korea (Registration Number: KCT0009946).

## Introduction

Temporomandibular disorder (TMD) is a musculoskeletal disorder that affects the temporomandibular joints (TMJ), masticatory muscles, and surrounding tissue structures [1]. As a critical joint in mastication and speech, the TMJ enables the essential movement of opening and closing the mouth, serving as the central axis of the lower jaw, while supporting the skull. Excessive force or overuse of the TMJ can increase fatigue, leading to joint pain, possible deformation, and functional decline, which can subsequently result in systemic complications.

An analysis of the National Health Insurance Service (NHIS) data from 2014 to 2023 in Korea revealed that the number of patients diagnosed with "temporomandibular joint disorders" (primary disease code K07.6) increased from 338,000 to 542,000 in the last 10 years, indicating a growing burden on healthcare resources. The total medical expenses for TMD increased from 37.8 billion KRW in 2014 to 109.8 billion KRW in 2023, reflecting an average annual increase of 13.7% [2]. To provide a comprehensive and standardized diagnosis and evaluation of TMD, the Diagnostic Criteria for Temporomandibular Disorders (DC/TMD) were introduced in 2014 as an updated framework of the earlier Research Diagnostic Criteria for Temporomandibular Disorders established in 1992 [3]. The symptoms of TMD are diverse and include discomfort and pain in the jaw joint, difficulty with speech and chewing, tinnitus, and clicking or popping sounds during jaw movements. When left untreated, persistent and progressive symptoms can deteriorate quality of life and potentially exacerbate psychiatric conditions such as depression, chronic stress, and anxiety [4].

Patients with TMD often experience improvement through a combination of noninvasive therapies, including education, cognitive behavioral therapy, pharmacological treatment, physical therapy, and occlusal devices. The initial treatment typically involves nonsteroidal anti-inflammatory drugs (NSAIDs) and muscle relaxants, with benzodiazepines or antidepressants considered for managing chronic conditions [5]. Despite the conventional treatments mentioned above, limited therapeutic effects can result in persistent pain and disabilities, negatively affecting patients’ quality of life, and occasionally leading to adverse effects [6,7]. Given these limitations, complementary Traditional Korean Medicine (TKM), mainly acupuncture, electroacupuncture, and Chuna manual therapy, has been used for the treatment of TMD.

Thread embedding acupuncture (TEA) is a promising alternative treatment for the treatment of TMJ disorders. TEA is a unique acupuncture method that involves the insertion of biodegradable threads into acupoints, providing continuous stimulation and tissue remodeling to relieve pain and improve function [8]. Several randomized controlled trials have reported positive outcomes including significant pain reduction, enhanced masticatory function, and increased maximum interincisal opening [9–11].

Previously, we conducted a pilot clinical trial to evaluate the efficacy, safety, and cost-effectiveness of TEA in the treatment of TMD [12]. Building on the findings of the pilot study, this confirmatory clinical trial was designed using an optimized protocol based on sample size calculations from the initial research. This study primarily aimed to provide definitive evidence for the efficacy of TEA in reducing pain intensity in patients with chronic TMD compared with physical therapy. The secondary objectives were to evaluate the safety, functional outcomes, improvement in quality of life, and cost-effectiveness of TEA for treating chronic TMD.

## Methods

### Study design

This study is a single-center randomized controlled trial (RCT) with two parallel groups: TEA and usual care. This trial will be conducted at Kyung Hee University Oriental Medicine Hospital at Gangdong and will adhere to the principles outlined in the Declaration of Helsinki. The study protocol was approved by the Institutional Review Board (IRB) of Kyung Hee University Oriental Hospital at Gangdong (Approval Number: KHNMCOH IRB 2024-08-002) and registered with the Clinical Research Information Service of the Republic of Korea (Registration Number: KCT0009946). The clinical procedures for TEA were developed according to the updated Standards for Reporting Interventions in Clinical Trials of Acupuncture (STRICTA) guidelines [13]. The protocol was based on the Standard Protocol Items: Recommendations for Interventional Trials (SPIRIT) [14]. This trial began with patient registration on October 25, 2024, and is currently ongoing.

### Participants

A total of 76 participants, aged 19–70 years diagnosed with chronic TMJ disorders based on the DC/TMD criteria will be recruited. After obtaining informed consent, the participants were screened using inclusion and exclusion criteria. Eligible participants were randomly assigned to either the experimental group (TEA treatment, n=38) or the control group (usual care, n=38).

#### Inclusion criteria

Participants who meet the following inclusion criteria will be included in this study:

1. Regional pain in the TMJ and surrounding areas (temple, inner ear, and front of the ear) persisting for more than three months, related to jaw movement, function, or parafunction, according to the DC/TMD.
2. Diagnosis of a pain disorder confirmed through palpation of the masticatory muscles or TMJ by the examiner, according to the DC/TMD criteria.
3. Average pain intensity of ≥40 mm on a 100 mm Visual Analogue scale (VAS) over the past week, with the more severe side assessed in cases of bilateral pain.
4. Adults aged 19–70 who have received a full explanation of the study and provided signed informed consent.

#### Exclusion criteria

Participants who meet any of the following criteria will be excluded from the study.

1. History of TEA or injection treatment in the facial region within the past three months.
2. Changes in the use of pain-related medications, including NSAIDs, within the past month.
3. Previous surgery involving the TMJ.
4. Multiple pain disorders (e.g., rheumatoid arthritis) or neurological conditions (e.g., trigeminal neuralgia and herpes zoster) that can influence TMJ pain.
5. Skin conditions or blood coagulation disorders (e.g., inflammation, wounds, and anticoagulant use) that contraindicate thread embedding acupuncture.
6. Pregnancy or planned pregnancy
7. Mental disorders or communication difficulties that would prevent adherence to clinical trial protocols.

#### Procedures

A total of 76 participants with TMJ pain disorder persisting more than three months were recruited from the Kyung Hee University Oriental Medicine Hospital at Gangdong. Detailed information regarding the study will be provided to all potential participants beforehand, and only those who agree to participate and sign an informed consent form will undergo eligibility screening. Those who meet the inclusion and exclusion criteria were randomly assigned to the TEA or usual care groups. The study will consist of a screening period, six treatment sessions (Week 0–5) over six weeks and two additional follow-up sessions (Week 6 and 10), and will be conducted according to the designated scheduled appointment. (Fig 1)

**Figure 1.**
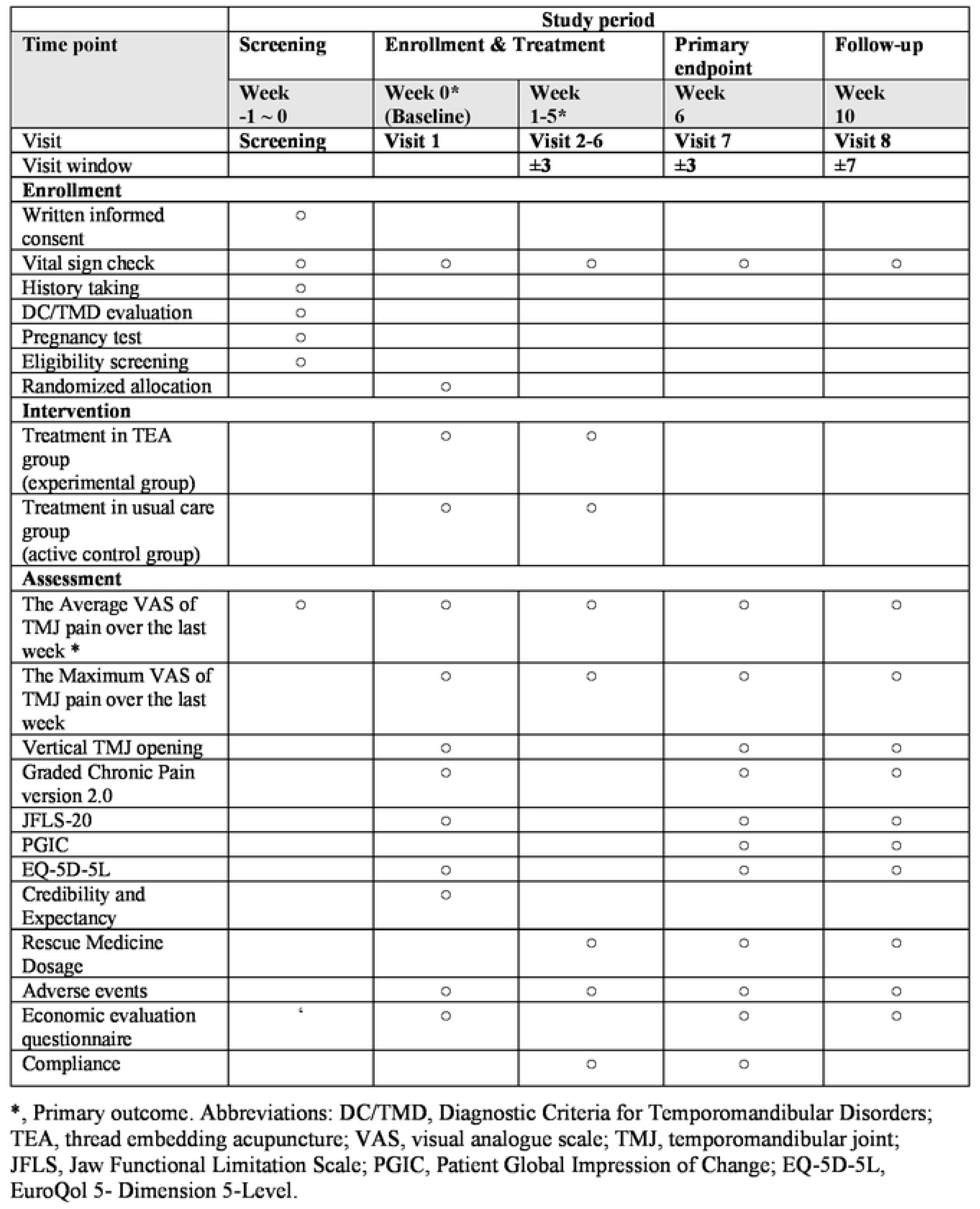
SPIRIT schedule of enrollment, interventions, and assessments.

### Interventions

Participants in the experimental group will receive TEA weekly for six weeks. The usual care group will receive physical therapy consisting of transcutaneous electrical nerve stimulation (TENS) and infrared (IR) therapy. (Fig 2)

**Figure 2.**
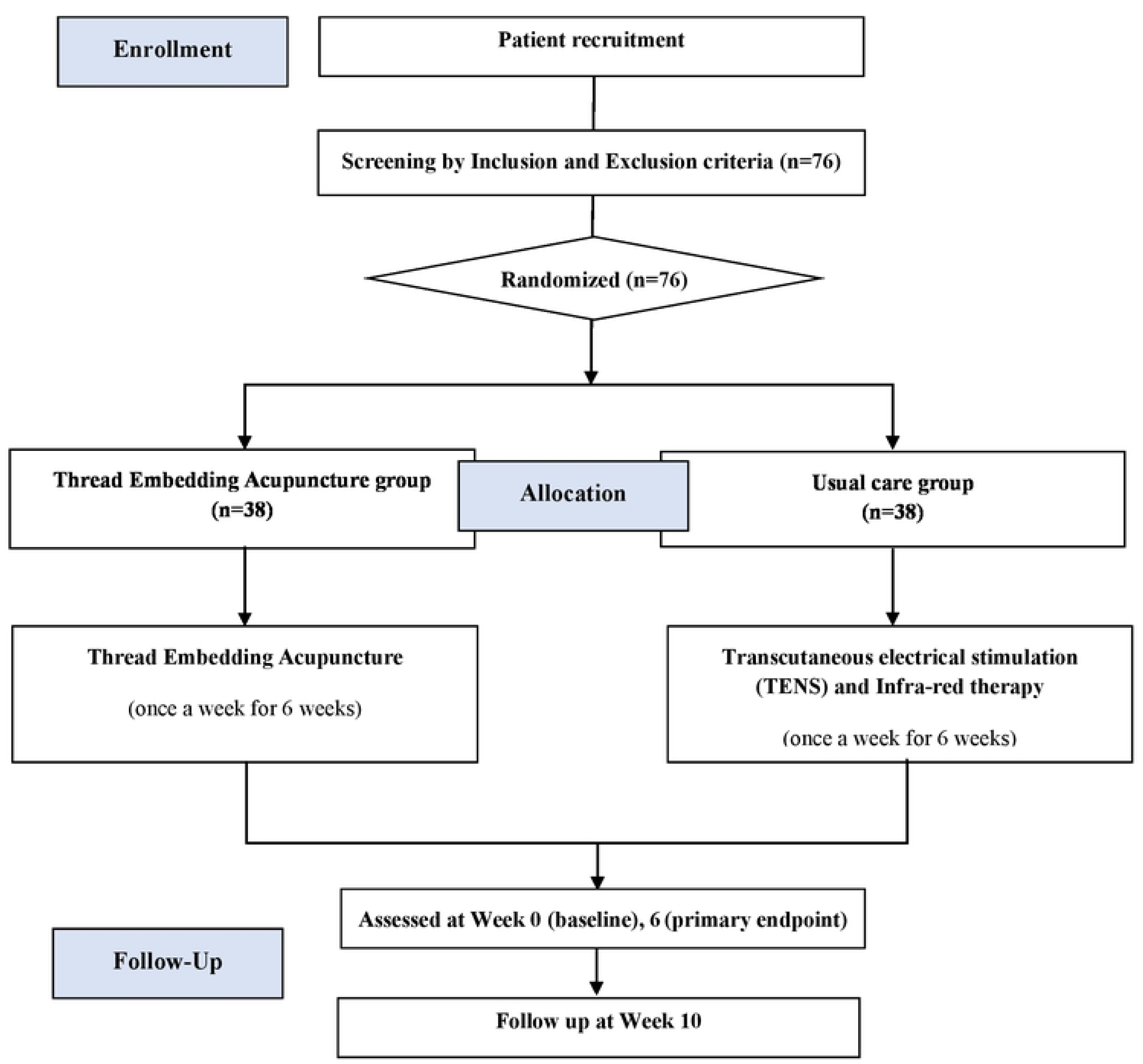
Study flow chart.

#### TEA group

The TEA group will be treated with TEA once week for six weeks. A total of 16 TEA (Dongbang Medical, Sungnam, South Korea) 29G x 25 mm needles with 30 mm threads were inserted into eight bilateral predefined acupoints. (Fig 3) The TEA procedure will be administered bilaterally to the TMJ, irrespective of the side of pain. Further details regarding the treatment group interventions, including acupoints and insertion techniques, are provided in the STRICTA checklist (Table 1) and illustrated in Fig 3A.

1. Daying (ST5) → Jiache (ST6): Transverse insertion along the line between ST5 and ST6 in the superficial fascia. (main line)
2. Daying (ST5) → Jiache (ST6) Superior Line: Transverse insertion 10 mm above the main line.
3. Jiache (ST6) → Xiaguan (ST7): Transverse insertion from ST6 to ST7 at the superficial fascia.
4. Jiaosun (TE20) → Shuaigu (GB8): Transverse insertion from TE20 to GB8 at the superficial fascia.
5. Shangguan (GB3) → Qubin (GB7): Transverse insertion from GB3 to GB7 in the superficial fascia.
6. Tianyou (TE16) → Yifeng (TE17): Transverse insertion from TE16 to TE17 in the superficial fascia.
7. Jianjing (GB21) → Jugu (LI16): Transverse insertion from GB21 to LI16 at the superficial fascia.
8. Xiaguan (ST7): Oblique insertion towards the space between the zygomatic arch and mandibular notch.

**Figure 3.**
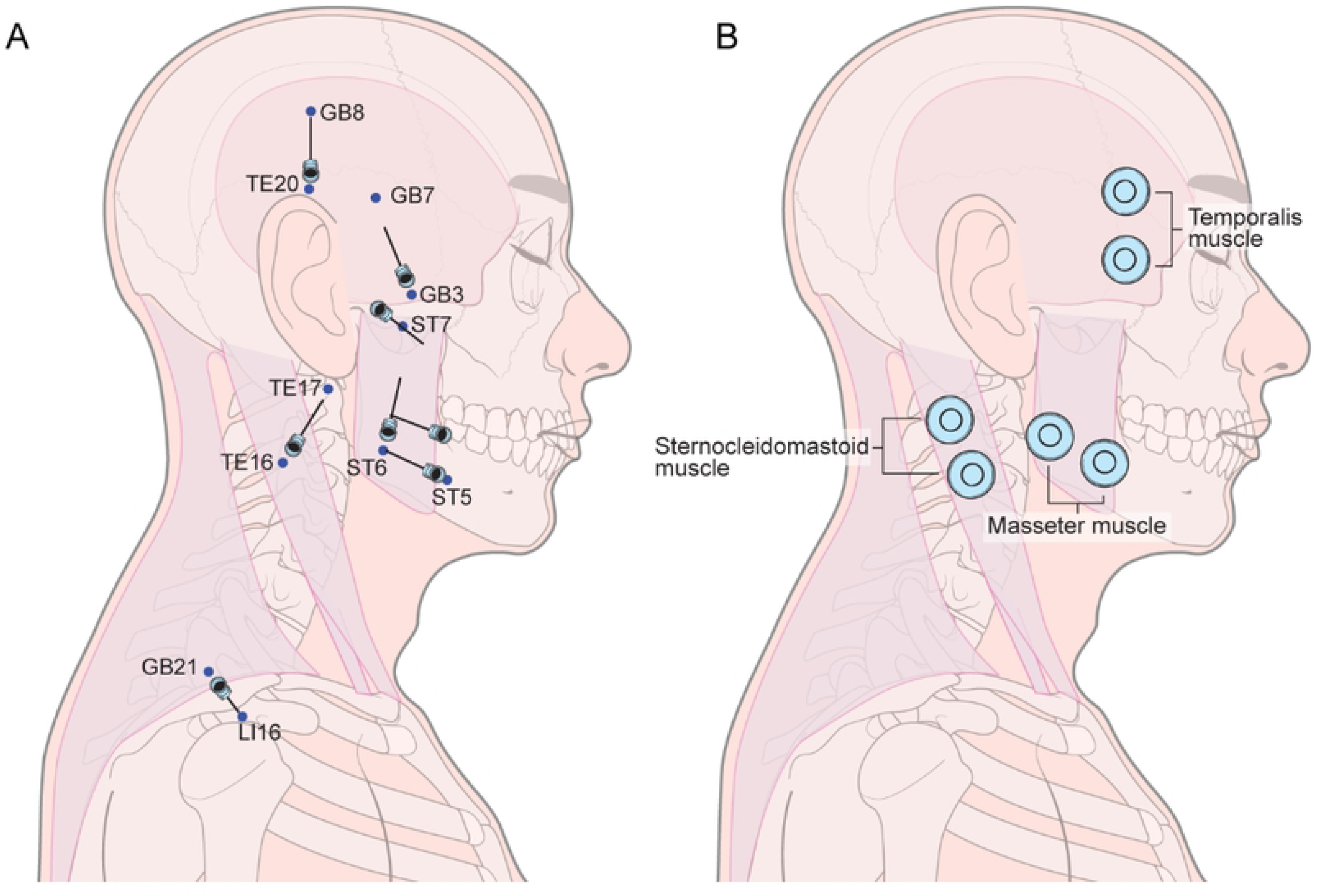
Location of intervention. (A) The designated acupoints and insertion directions for the thread embedding acupuncture group are illustrated. (B) The specific areas targeted for transcutaneous electrical nerve stimulation in the usual care group are depicted, where three pairs of electrodes are applied. Infrared therapy will be simultaneously administered to these same regions.

**Table 1.**
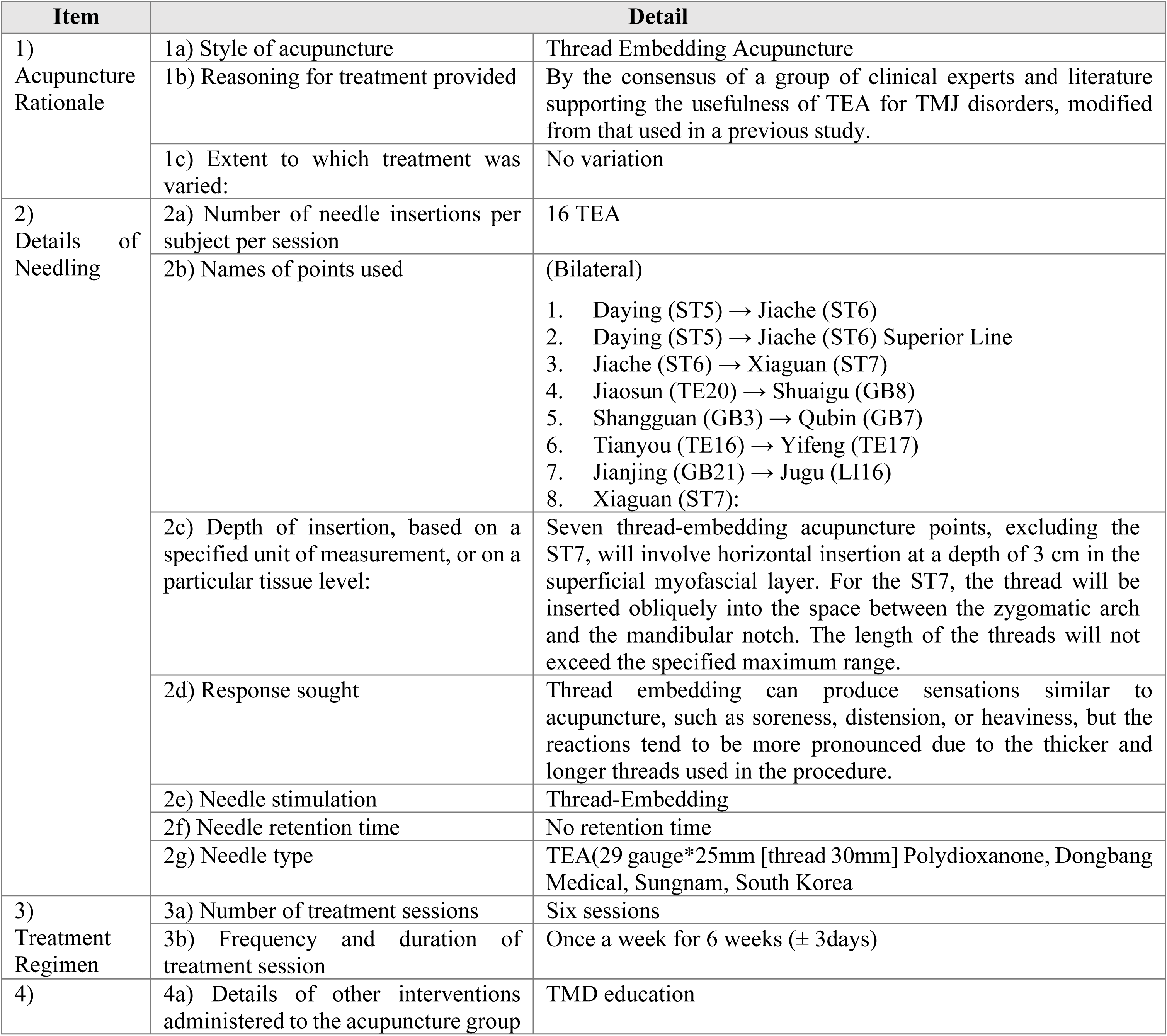
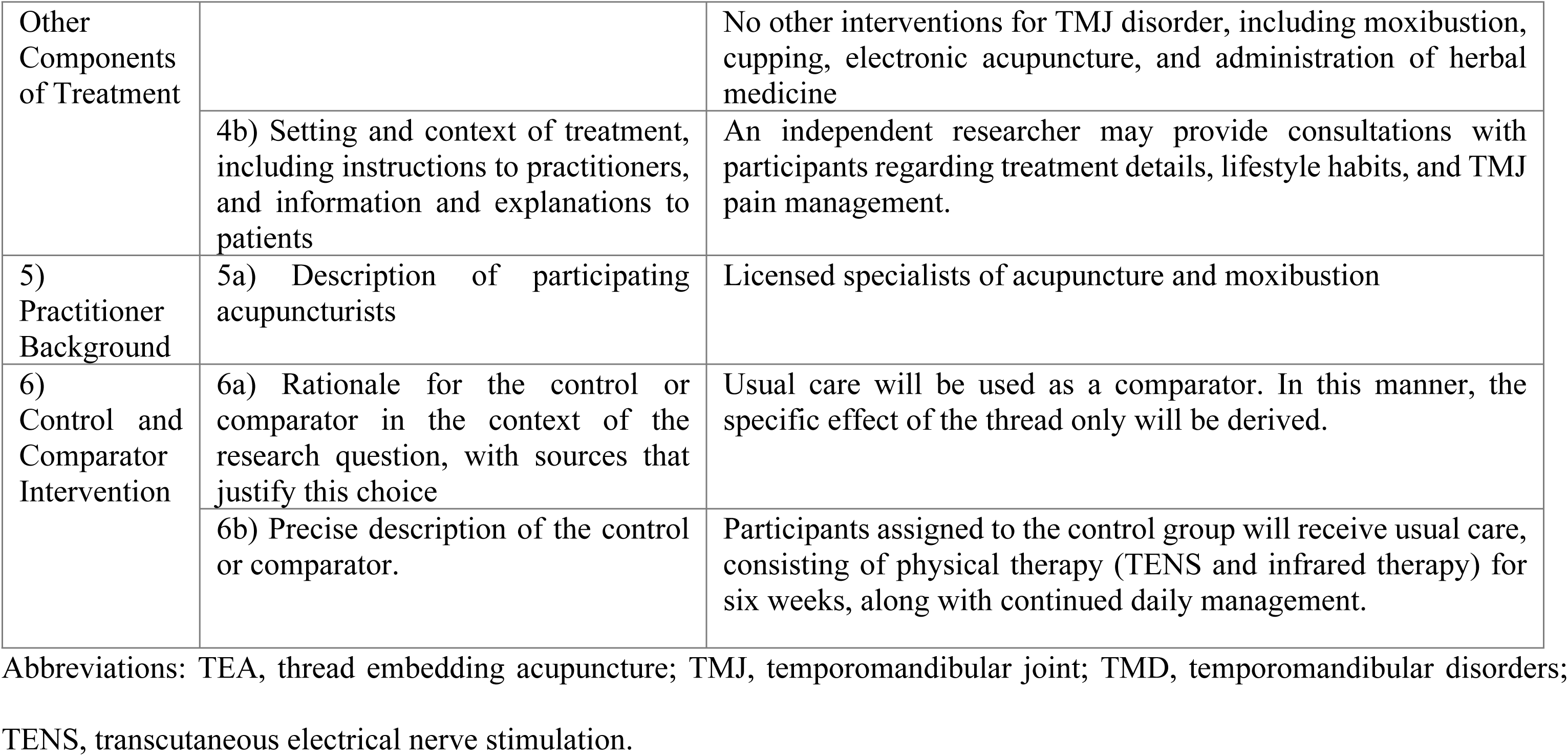
Checklists for the TEA treatment According to STRICTA (Standard Reporting In Controlled Trials of Acupuncture) Recommendations.

#### Usual care group

The usual care group will receive weekly physical therapy for six weeks, involving TENS (SPMI-330®, Stratek, Anyang, South Korea) set at alternating currents of 2 Hz and 100 Hz every 5 s for 15 min, combined with IR therapy (IR-300®, Daekyung, Pocheon, South Korea). TENS pads were attached bilaterally to the temporalis, masseter, and sternocleidomastoid muscles, with a pair of electrodes placed on each muscle, resulting in six stimulation sites. Stimulation intensity was adjusted to the level at which the participant could perceive electrical stimulation without experiencing pain or discomfort. Simultaneously, IR therapy will be applied at an intensity of up to 250 W, while maintaining a minimum distance of 20–30 cm from the participant.

#### TMD education

TMD education will be provided to all participants, regardless of the group assignment. This education program will include materials on the causes of TMD, prevention, management, and self-corrective stretching exercises.

#### Rescue medication and concurrent treatment

Throughout the study period, acetaminophen (e.g., Tylenol, Acetaminophen tablets) will be permitted as a rescue medication for all participants. Participants will be instructed to take 1–2 tablets of 500 mg acetaminophen per dose at a minimum interval of 4–6 h only when pain is intolerable, with a maximum daily dosage of eight tablets. The use of these medications was self-reported and documented. A total of 30 tablets were provided during weeks 1–4, and 50 tablets during week 5. At each subsequent visit, the remaining tablets were counted to verify adherence and usage.

### Outcome measures

The following assessments will be performed at baseline (Week 0), one week after treatment termination (Week 6), and five weeks after treatment termination (Week 10). The primary outcome will be the average pain intensity measured using a 100 mm VAS. Secondary outcomes will include maximum pain intensity (100 mm VAS), vertical TMJ opening, Graded Chronic Pain 2.0 (GCP 2.0), Jaw Functional Limitation Scale-20 (JFLS-20), Patient Global Impression of Change (PGIC), EuroQol 5-Dimension 5-level (EQ-5D-5L), adverse events, and medical costs for economic evaluation. These assessments were used to evaluate the efficacy, safety, and cost-effectiveness of TEA in the treatment of chronic TMD.

#### Primary outcome

- **Average pain VAS.** The average intensity of TMJ pain over the last week will be evaluated using the VAS score, which consists of a horizontal 100 mm line, where one end represents "no pain" and the other end represents "the most severe pain imaginable.” Participants will be asked to mark a single point reflecting their average TMJ pain over the past week along this line during the screening period, weeks 0–5 (treatment), 6 (primary endpoint), and 10 (follow-up period). For patients experiencing unilateral pain or differing pain intensities in the bilateral TMJs, the side with more severe symptoms will be used as the reference for recording.

#### Secondary outcomes

- **Maximum pain VAS.** Participants will be asked to mark a point reflecting the maximum TMJ pain over the previous week on a 100-mm linear scale. The maximum pain VAS scores will be assessed at Week 0–5, 6, and 10.
- **Vertical TMJ opening.** The vertical range of mouth opening will be measured in accordance with the DC/TMD and classified into “pain-free opening” and “maximum unassisted opening.” Measurements will be performed using the TheraBite® (Atos Medical, Horby, Sweden), a range of motion scale. The vertical TMJ opening was assessed at weeks 0, 6, and 10. *Pain-Free Opening* was measured after instructing the patient to open the mouth as wide as possible without feeling any pain or worsening current pain. *Maximum Unassisted Opening* was measured after instructing the patients to open their mouth as wide as possible, even if it caused pain.
- **GCP 2.0.** The GCP 2.0, from the DC/TMD, evaluates the characteristic pain intensity (CPI), pain duration, degree of interference in daily activities, and duration of interference. The CPI was calculated as the average score of the three pain intensity items. The total disability points will be determined using the grading criteria provided by the DC/TMD, based on the average scores for the duration of interference with daily activities and the degree of interference in daily life. GCP 2.0, will be assessed at weeks 0, 6, and 10.
- **JFLS-20.** The JFLS-20 evaluates jaw function over the past month, focusing on chewing, jaw movement, and functions related to communication and emotional expression. It consists of 20 items with responses scored on a scale from 0 (no restriction) to 10 (severe restriction). The Korean version of the JFLS-20 used in this study was officially provided by the International Network for Orofacial Pain and Related Disorders Methodology (INFORM). JFLS-20 will be assessed at weeks 0, 6, and 10. PGIC. The PGIC is a subjective method for patients to evaluate the degree of improvement on a 7-point scale: 1 (very much improved), 2 (much improved), 3 (minimally improved), 4 (no change), 5 (minimally worse), 6 (much worse), or 7 (very much worse). The PGIC will be assessed at weeks 6 and 10.
- **EQ-5D-5L.** The EQ-5D-5L is one of the most widely used indirect methods for evaluating health states and consists of five items that assess the severity of issues related to mobility, self-care, usual activities, pain/discomfort, and anxiety/depression. Each item is assigned a specific weight depending on its severity level. These weights, along with constants, were incorporated into an equation to calculate the preference score. EQ-5D-5L will be assessed at weeks 0, 6, and 10.
- **Credibility and Expectancy.** Participants’ expectations regarding treatment will be assessed using a 9-point Likert scale. During the initial visit (week 0), the participants responded to the question, “To what extent do you believe TEA or usual care therapy will alleviate your symptoms?” by selecting scores ranging from 1 (not at all) to 5 (somewhat) to 9 (very much).
- **Rescue Medication consumption**. The dose of rescue medication administered during the study period was recorded at each visit. The types and doses of medications taken during the study period (prescribed for current medical conditions) will be monitored using questionnaires administered during participant visits. In addition to medication, the frequency of other treatments such as physical therapy or injection therapy, was also recorded.

#### Adverse events (AE)

Detailed data related to adverse effects (AE) will be collected at every visit, including symptom characteristics, dates of onset and resolution, severity and occurrence of serious adverse events (SAE), their potential relationship to clinical trial procedures, actions taken in response to trial-related procedures, interventions for AEs, and outcomes associated with these AEs.

Collected safety data will be appropriately summarized, with all SAEs described narratively, and assessed by comparing the frequency of AEs between groups. AEs will also be classified based on the Spilker classification system as mild (no significant interference with normal activities or functions), moderate (significant interference with activities or requiring treatment), or severe (serious adverse reactions, lasting sequelae, and requiring intensive treatment). The causal relationship between each treatment method and reported AEs will be investigated using a six-point scale: 1 (definitely related), 2 (probably related), 3 (possibly related), 4 (probably not related), 5 (definitely not related), and 6 (unknown).

#### Medical costs for economic evaluation

To measure the cost-effectiveness of the treatment for chronic TMD, a specifically developed structured questionnaire was used. Formal medical costs refer to the expenses incurred by utilizing healthcare services at medical institutions for the prevention, treatment, or rehabilitation of TMD. In contrast, informal medical costs include out-of-pocket expenditures for items such as health supplements and medical devices. The medical costs will be assessed at weeks 0, 6, and 10.

### Sample size

Using an empirical method for continuous variables and data from a previous pilot study, the mean difference (14.07) and standard deviation (20.225) of the primary efficacy outcome measure–the Physical Score of the Facial Disability Index–were used to calculate the required sample size. This calculation, using the following formula with an appropriate effect size, also supports the safety of TEA in clinical research [15].

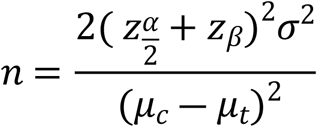

Considering the dropout rate of 10%, 76 participants were required. In a 1:1 ratio, 38 participants will be assigned to each of the TEA and usual care groups.

### Criteria for withdrawal and termination

In accordance with ethical principles protecting patient autonomy, participants have the right to withdraw from the study at any time and for any reason. The study may be discontinued in cases of consent withdrawal by the participant, the presence of a medical condition that endangers the participant’s safety, or the investigator’s determination that further participation is inappropriate for other reasons. I n the event of a severe adverse reaction, the investigator will permanently discontinue the clinical trial for the affected participant.

### Randomization and allocation concealment

Participants who signed the informed consent form and met the eligibility criteria were assigned to groups based on a pre-generated randomization table. Each participant will receive a unique identification code (Randomized Number) in the order of recruitment and treatments will be administered based on their assigned group. The randomization table will be generated by an independent researcher not involved in the clinical trial using the block randomization method in Excel 2019 (Microsoft Corporation, Redmond, WA, USA), with the block size concealed from those conducting recruitment and assignments [16].

### Blinding

The randomization table will be managed by an independent researcher who is not involved in the treatment or evaluation processes. The participants were assigned to groups by sequentially opening sealed numbered envelopes based on the randomization table. These envelopes will be managed by an independent envelope administrator unrelated to the study. The randomization process will remain concealed until allocation and will be irreversible thereafter.

Due to the nature of the intervention, blinding of the practitioner and participants is not feasible; therefore, only assessor blinding was implemented. A trained blinded assessor, uninvolved in the treatment, will conduct evaluations in a separate area without knowledge of the group assignments.

Unblinding will be considered on a case-by-case basis only in serious medical emergencies and must be approved by the principal investigator. All unblinded procedures were thoroughly documented.

### Statistical methods

The efficacy assessment data from the participants will be analyzed to compare differences within and between groups. The Full Analysis Set (FAS), including all participants who received at least one treatment, is used for efficacy evaluation, and missing data are handled using the last observation carried forward (LOCF) method. Within-group differences before and after treatment will be analyzed using paired t-tests, whereas between-group differences in pre- and post-treatment changes are assessed using independent t-tests. Interaction effects between time and group will be evaluated using repeated-measures analysis of variance (ANOVA). All analyses are conducted at a significance level of 0.05, using the SAS version 9.1.3 statistical package (SAS Institute, Cary, NC, USA), with significance set at p<0.05.

The economic evaluation adopts a healthcare system perspective, incorporating formal and informal medical costs based on data from the Health Insurance Review and Assessment Service and the National Health Insurance Service. Costs and quality of life, measured using the EQ-5D-5L, will be analyzed following the intention-to-treat (ITT) principle from baseline to the study’s conclusion, with multiple imputations for missing data and extrapolation methods applied if needed.

Cost-effectiveness will be evaluated using the incremental cost-effectiveness ratio (ICER) with quality-adjusted life years (QALY) derived from EQ-5D-5L data, utilizing the area under the curve method. Bootstrapping generated cost-effectiveness planes and acceptability curves with a willingness-to-pay threshold of 30.5 million KRW (USD 25,000). Sensitivity analyses account for clinical validity and uncertainty. To ensure safety, the incidences of all AEs reported in this study was monitored.

### Data collection and management

Participants and clinical trial information will be documented in the electronic medical record (EMR) system of the medical institution. Data collection, management procedures, and information on blinding will be provided only to designated researchers who will handle and crosscheck the trial data. Evaluation records completed by patients and assessors will be kept confidential and maintained as supporting evidence, recorded in both the EMR and paper, and subsequently entered into an electronic case report form (eCRF) provided by MyTrial (Bethesda Soft Co., Ltd., Seoul, Korea). Researchers are assigned role-based permissions to access, modify, and input data within the eCRF system, with all activities logged for accountability.

### Quality control

The study procedures and documentation will be periodically monitored according to the Korean Good Clinical Practice guidelines.

## Discussion

This randomized controlled trial is designed to evaluate the efficacy, safety, and cost-effectiveness of TEA compared with usual care for chronic TMD. Participants will be selected according to the DC/TMD and will receive either TEA or physical therapy consisting of TENS and IR therapy.

Current primary treatment approaches for TMD aim to increase the range of motion of the TMJ, alleviate pain and inflammation in the joint and masticatory muscles, and prevent degenerative changes in joint tissues. These approaches typically involve a combination of pharmacological and non-pharmacological treatments. Non-pharmacological treatments commonly include patient education, self-management, occlusal stabilization devices, and physical therapy. Pharmacological treatments include the prescription of NSAIDs, antidepressants, and muscle relaxants [17]. Notably, TENS, used as a control intervention in this study, has been reported to clinically reduce pain intensity in individuals with temporomandibular disorders, supported by moderate-quality evidence [18]. To establish a robust control group, TENS was applied using alternating modes of low (2 Hz) and high (100 Hz) frequencies, which were expected to synergistically enhance analgesic effects via dynorphin and enkephalin activation [19].

Acupuncture has shown promising results in the management of TMD. Recent studies have reported that acupuncture can substantially reduce TMJ pain intensity and improvements in physical and emotional functions [20]. Targeting trigger points with acupuncture in the masticatory muscles, including the lateral pterygoid, medial pterygoid, and temporalis muscles, can effectively relieve muscle stiffness and contracture [21]. It modulates TMD-related pain by influencing the nervous system, reducing the expression of inflammatory receptors, and regulating the hypothalamic-pituitary-adrenal axis [22].

TEA is a novel approach for inserting biodegradable threads at specific points to provide sustained acupuncture stimulation. This method has been shown to be effective in the management of chronic musculoskeletal pain [23,24]. Polydioxanone, the thread material used in TEA, exerts physical and chemical effects by utilizing the tension of the thread itself to pull the tissue and promoting collagen formation. It stimulates fibroblast proliferation through the TGF-β signaling pathway, promoting collagen formation and achieving tissue remodeling [8]. TEA can relieve muscle tension, improves blood flow, and reduces inflammation by targeting the muscle and fascial layers. These features make it particularly effective for pain-related TMD conditions such as myalgia, myofascial pain, and arthralgia. In the same context, participants of this study were selected using the DC/TMD criteria 3 and 4, which focus on localized pain in the jaw, temples, ear, or front of the ear over the past 30 d, as well as pain in the masticatory muscles or TMJ, as confirmed by palpation.

The intervention methodology employed in this study was based on a survey investigating the current use of TEA for the treatment of TMD [25]. The frequency of interventions was determined based on survey responses, indicating that an average of 5.9 sessions was considered the minimum effective number of TEA treatments to achieve significant therapeutic effects against TMD. Key acupoints, such as Jiache (ST6) and Yifeng (TE17), were selected based on the survey results, considering their anatomical relevance to the TMJ and surrounding muscles. Building on a previous pilot study, TEA will be applied to the trapezius muscle in addition to the masseter, temporalis, and sternocleidomastoid muscles. Silveira et al. [26] demonstrated that cervical disorders can influence TMJ dysfunction, with greater pain in the trapezius and temporalis muscles associated with higher levels of TMD severity. In addition, the safety of TEA was referenced from a previous study, which reported that out of 89 cases of facial TEA procedures, 8 cases (9%) experienced adverse effects [27]. To ensure the safety of the participants, this study proactively profiled the safety and potential side effects of TEA in the facial area based on previous studies.

This RCT addressed the limitations of previous studies, such as small sample sizes and lack of comprehensive economic evaluations. Participants will be assessed using validated outcome measures, such as the VAS for pain intensity, JFLS-20 for functional limitations, and EQ-5D-5L for quality of life. These measures provide a holistic understanding of the effects of TEA on the multifaceted nature of TMD, extending beyond simple pain relief. Additionally, the study incorporated a cost-effectiveness analysis comparing TEA with physical therapy using ICER and QALYs. By addressing both the clinical and economic outcomes, these findings can inform healthcare policies and support the integration of TEA into standard TMD treatment protocols.

Despite its strengths, this study had certain limitations. The single-center design may restrict the generalizability of the findings, and the relatively short follow-up period may not capture the long-term outcomes or delayed adverse effects. Additionally, blinding of the practitioner and participants was not feasible because of the nature of the intervention. Future multicenter studies with extended follow-up periods are recommended to validate these findings and to explore the long-term safety and efficacy of TEA in TMD management.

## Conclusion

We designed this RCT to evaluate the efficacy, safety, and cost-effectiveness of TEA for the treatment of TMD. The findings of this study are expected to expand the clinical application of TEA as an effective and cost-effective treatment for TMD.

## Data Availability

All relevant data are within the manuscript and its Supporting Information files.

https://cris.nih.go.kr/cris/search/detailSearch.do?search_lang=E&focus=reset_12&search_page=L&pageSize=10&page=undefined&seq=28597&status=5&seq_group=28597

## Supporting information

S1 File. SPIRIT checklist

(DOC)

## Ethics statement

The study protocol was approved by the IRB of Kyung Hee University Oriental Medicine Hospital at Gangdong [IRB Number: KHNMCOH IRB 2024-08-002]. Written informed consent was obtained from all the participants. This study was conducted in compliance with the ethical guidelines of the Declaration of Helsinki.

## Acknowledgments

We would like to thank Editage (www.editage.co.kr) for editing and reviewing this manuscript for English language.

## Author Contributions

**Conceptualization:** Bonhyuk Goo, Sang-Soo Nam.

**Data curation:** Bonhyuk Goo, Jung-Hyun Kim.

**Funding Acquisition:** Sang-Soo Nam

**Formal analysis:** Jinkyung Park, Bonhyuk Goo.

**Investigation:** Jinkyung Park, Jaeho Song, Chanju Im, Jun-Gyu Park.

**Methodology:** Bonhyuk Goo, Sang-Soo Nam, Jung-Hyun Kim.

**Project Administration:** Bonhyuk Goo.

**Supervision:** Sang-Soo Nam

**Validation:** Jung-Hyun Kim, Jaeho Song, Chanju Im, Jun-Gyu Park.

**Writing-original draft:** Jinkyung Park

**Writing-review & editing:** Sang-Soo Nam, Bonhyuk Goo, Jung-Hyun Kim, Jaeho Song, Chanju Im, Jun-Gyu Park.

## References

1. Valesan LF, Da-Cas CD, Réus JC, Denardin ACS, Garanhani RR, Bonotto D, et al. Prevalence of temporomandibular joint disorders: a systematic review and meta-analysis. Clin Oral Investig. 2021;25: 441–453. doi:10.1007/s00784-020-03710-w

2. Disease Subdivision (4-tier classification disease) Statistics. The number of patients by year when K076 is entered in the disease code inquiry. 2023 [cited 23 Jan 2025]. Available: https://opendata.hira.or.kr/op/opc/olap4thDsInfoTab1.do

3. Schiffman E, Ohrbach R, Truelove E, Look J, Anderson G, Goulet J-P, et al. Diagnostic Criteria for Temporomandibular Disorders (DC/TMD) for Clinical and Research Applications: recommendations of the International RDC/TMD Consortium Network* and Orofacial Pain Special Interest Group†. J Oral Facial Pain Headache. 2014;28: 6–27. doi:10.11607/jop.1151

4. Warzocha J, Gadomska-Krasny J, Mrowiec J. Etiologic Factors of Temporomandibular Disorders: A Systematic Review of Literature Containing Diagnostic Criteria for Temporomandibular Disorders (DC/TMD) and Research Diagnostic Criteria for Temporomandibular Disorders (RDC/TMD) from 2018 to 2022. Healthcare (Basel). 2024;12. doi:10.3390/healthcare12050575

5. Gauer RL, Semidey MJ. Diagnosis and treatment of temporomandibular disorders. Am Fam Physician. 2015;91: 378–86.

6. List T, Axelsson S. Management of TMD: evidence from systematic reviews and meta-analyses. J Oral Rehabil. 2010;37: 430–51. doi:10.1111/j.1365-2842.2010.02089.x

7. Fischoff D, Spivakovsky S. Are pharmacological treatments for oro-facial pain effective? Evid Based Dent. 2018;19: 28–29. doi:10.1038/sj.ebd.6401294

8. Ha YI, Kim JH, Park ES. Histological and molecular biological analysis on the reaction of absorbable thread; Polydioxanone and polycaprolactone in rat model. J Cosmet Dermatol. 2022;21: 2774–2782. doi:10.1111/jocd.14587

9. Dietrich L, Rodrigues IVS, Assis Costa MDM de, Carvalho RF, Silva GR da. Acupuncture in Temporomandibular Disorders Painful Symptomatology: An Evidence-Based Case Report. Eur J Dent. 2020;14: 692–696. doi:10.1055/s-0040-1716631

10. Jung A, Shin B-C, Lee MS, Sim H, Ernst E. Acupuncture for treating temporomandibular joint disorders: a systematic review and meta-analysis of randomized, sham-controlled trials. J Dent. 2011;39: 341–50. doi:10.1016/j.jdent.2011.02.006

11. Cho S-H, Whang W-W. Acupuncture for temporomandibular disorders: a systematic review. J Orofac Pain. 2010;24: 152–62.

12. Kim J-H, Kang D, Kim K-W, Nam S-S, Goo B. Thread Embedding Acupuncture for Temporomandibular Disorder: Protocol for a Pilot Randomized Controlled Trial. J Pain Res. 2022;15: 3197–3207. doi:10.2147/JPR.S383965

13. MacPherson H, Altman DG, Hammerschlag R, Youping L, Taixiang W, White A, et al. Revised STandards for Reporting Interventions in Clinical Trials of Acupuncture (STRICTA): extending the CONSORT statement. PLoS Med. 2010;7: e1000261. doi:10.1371/journal.pmed.1000261

14. Chan A-W, Tetzlaff JM, Altman DG, Laupacis A, Gøtzsche PC, Krleža-Jerić K, et al. SPIRIT 2013 statement: defining standard protocol items for clinical trials. Ann Intern Med. 2013;158: 200–7. doi:10.7326/0003-4819-158-3-201302050-00583

15. Whitehead AL, Julious SA, Cooper CL, Campbell MJ. Estimating the sample size for a pilot randomised trial to minimise the overall trial sample size for the external pilot and main trial for a continuous outcome variable. Stat Methods Med Res. 2016;25: 1057–73. doi:10.1177/0962280215588241

16. Kang H. Random allocation and dynamic allocation randomization. Anesth Pain Med. 2017;12: 201–212. doi:10.17085/apm.2017.12.3.201&domain=pdf&date_stamp=2017-07-25

17. Butts R, Dunning J, Pavkovich R, Mettille J, Mourad F. Conservative management of temporomandibular dysfunction: A literature review with implications for clinical practice guidelines (Narrative review part 2). J Bodyw Mov Ther. 2017;21: 541–548. doi:10.1016/j.jbmt.2017.05.021

18. Serrano-Muñoz D, Beltran-Alacreu H, Martín-Caro Álvarez D, Fernández-Pérez JJ, Aceituno-Gómez J, Arroyo-Fernández R, et al. Effectiveness of Different Electrical Stimulation Modalities for Pain and Masticatory Function in Temporomandibular Disorders: A Systematic Review and Meta-Analysis. J Pain. 2023;24: 946–956. doi:10.1016/j.jpain.2023.01.016

19. Law P, Cheing G. Optimal stimulation frequency of transcutaneous electrical nerve stimulation on people with knee osteoarthritis. J Rehabil Med. 2004;36: 220–225. doi:10.1080/16501970410029834

20. Liu L, Chen Q, Lyu T, Zhao L, Miao Q, Liu Y, et al. Effect of acupuncture for temporomandibular disorders: a randomized clinical trial. QJM. 2024;117: 647–656. doi:10.1093/qjmed/hcae094

21. Simma I, Simma L, Fleckenstein J. Muscular diagnostics and the feasibility of microsystem acupuncture as a potential adjunct in the treatment of painful temporomandibular disorders: results of a retrospective cohort study. Acupuncture in medicine: journal of the British Medical Acupuncture Society. 2018;36: 415–421. doi:10.1136/acupmed-2017-011492

22. Serritella E, Colombo V, Özcan M, Galluccio G, Di Paolo C. Acupuncture and Traditional Chinese Medicine in the Management of Orofacial Pain and Temporomandibular Disorders: a Narrative Review. Curr Oral Health Rep. 2024;11: 59–67. doi:10.1007/s40496-023-00359-8

23. Lee JH, Yang TJ, Lee DG, Lee OJ, Wei TS. The Effect of Needle-embedding Therapy on Osteoarthritis of the Knee Combined with Korean Medical Treatment: Report of Five Cases ※. The Acupuncture. 2014;31: 195–204. doi:10.13045/acupunct.2014066

24. Lim SS, Sung HJ, Lee CK, Choi HY, Roh J Du, Lee EY. The Effect of Thread Embedding Acupuncture on Lumbar Herniated Intervertebral Disc Patients: A Retrospective Study ※. The Acupuncture. 2016;33: 39–47. doi:10.13045/acupunct.2016053

25. Yu SH, Kang J, Seo S, Seo J, Kim S, Lim J-H, et al. Clinical Use of Thread Embedding Acupuncture for Temporomandibular Joint Disorder: A Web-Based Survey. Journal of Korean Medicine Rehabilitation. 2023;33: 149–160. doi:10.18325/jkmr.2023.33.3.149

26. Silveira A, Gadotti IC, Armijo-Olivo S, Biasotto-Gonzalez DA, Magee D. Jaw dysfunction is associated with neck disability and muscle tenderness in subjects with and without chronic temporomandibular disorders. Biomed Res Int. 2015;2015: 512792. doi:10.1155/2015/512792

27. Ha S, Lee S, Goo B, Kim E, Kwon O, Nam SS, et al. Safety of Thread-Embedding Acupuncture: A Multicenter, Prospective, Observational Pilot Study. Healthcare (Switzerland). 2024;12. doi:10.3390/healthcare12232396

